# Whole-exome and Whole-genome Sequencing of 1097 Individuals with Type 1 Diabetes Reveals Novel Genes for Diabetic Kidney Disease

**DOI:** 10.1101/2023.11.13.23298447

**Authors:** Jani K Haukka, Anni A Antikainen, Erkka Valo, Anna Syreeni, Emma H Dahlström, Bridget M Lin, Nora Franceschini, Valma Harjutsalo, Per-Henrik Groop, Niina Sandholm, FinnDiane Study Group

**Affiliations:** Folkhälsan Institute of Genetics, Folkhälsan Research Center, Helsinki; Department of Nephrology, University of Helsinki and Helsinki University Hospital, Helsinki, Finland; Research Program for Clinical and Molecular Metabolism, Faculty of Medicine, University of Helsinki, Helsinki, Finland; Department of Epidemiology, University of North Carolina, Chapel Hill, NC, United States; Department of Diabetes, Central Clinical School, Monash University, Melbourne, Victoria, Australia

## Abstract

**Background and hypothesis:** Diabetic kidney disease (DKD) is a severe diabetic complication affecting one third of individuals with type 1 diabetes. Although several genes and common variants have been associated with DKD, much of the predicted inheritance remain unexplained. Here, we performed next-generation sequencing to assess whether low-frequency variants — single or aggregated — contribute to the missing heritability in DKD.

**Methods:** We performed whole-exome sequencing (WES) of 498 individuals and whole-genome sequencing (WGS) of 599 individuals with type 1 diabetes. After quality control, we had next-generation sequencing data available for altogether 1064 individuals, of whom 546 had developed either severe albuminuria or end-stage kidney disease, and 528 had retained normal albumin excretion despite a long duration of type 1 diabetes. Single variants and gene aggregate tests were performed separately for WES and WGS data and combined with meta-analysis. Furthermore, we performed genome-wide aggregate analyses on genomic windows (sliding-window), promoters, and enhancers with the WGS data set.

**Results:** In single variant meta-analysis, no variant reached genome-wide significance, but a suggestively associated *THAP7* rs369250 variant (*P*=1.50×10^-5^) was replicated in the FinnGen general population GWAS data for chronic kidney disease (CKD) and DKD phenotypes. Gene-aggregate meta-analysis identified suggestive evidence (*P*<4.0×10^-4^) at four genes for DKD, of which *NAT16* and *LTA* (TNB-β) replicated in FinnGen. Of the intergenic regions suggestively associated with DKD, the enhancer on chromosome 18q12.3 (*P*=3.94×10^-5^) showed interaction with the *METTL4* gene; the lead variant was replicated, and predicted to alter Mafb binding.

**Conclusions:** Our sequencing-based meta-analysis revealed multiple genes, variants and regulatory regions suggestively associated with DKD. However, as no variant or gene reached genome-wide significance, further studies are needed to validate the findings.

**What was known:**

- Genetics is an important factor in the development and progression of diabetic kidney disease (DKD) in individuals with type 1 diabetes.
- Previously identified genetic associations have mostly been common variants as they originated from GWAS studies. Based on inheritance estimates, the current findings only explain a fraction of the predicted disease risk.

**This study adds:**

- Our study with 1097 sequenced individuals with type 1 diabetes is to date one of the largest sequencing studies on DKD in type 1 diabetes.
- The study reveals several suggestive variants, genes and intergenic regulatory regions associated with DKD. Low-frequency protein-altering variants inside *NAT16* and *LTA* (encoding for TNF-β), and chromosome 18q12.3 enhancer variant linking to *METTL4* were also replicated in FinnGen kidney disease phenotypes.

**Potential impact:**

- The results suggest novel genes that may be important for the onset and development of serious DKD in individuals with type 1 diabetes. In addition to revealing novel biological mechanisms leading to DKD, they may reveal novel treatment targets for DKD. However, further validation and functional studies are still needed.

## INTRODUCTION

Type 1 diabetes (T1D) is an autoimmune disease caused by the destruction of the insulin-secreting beta cells in the islets of Langerhans in the pancreas. Long-term insulin irregularity leads to complications in several organs for a large proportion of individuals with T1D [1]. In particular, prolonged hyperglycaemia leads to a decline of kidney function and diabetic kidney disease (DKD) for approximately 30% of individuals with T1D [1, 2]. In the Western world, DKD is the most common cause of kidney failure, which can be treated only with dialysis or kidney transplantation [3]. In addition, DKD predisposes the individuals to cardiovascular disease (CVD), and already early-stage DKD (moderate albuminuria) elevates the risk of myocardial infarction and stroke two to three-fold [4, 5].

Both genetic and environmental factors affect the occurrence of T1D and its complications. Heritability estimates from genome-wide association studies (GWAS) suggest that genetic factors explain approximately one-third of the DKD risk [6, 7]. Microarray-based chips used in GWASs include hundreds of thousands of common human-variable loci, excellent for the study of common variants having modest effects on the disease risk [8, 9]. Indeed, GWASs have shed light upon DKD mechanisms, but these earlier findings explain only a minority of the predicted genetic risk of DKD [10-13]. Our recent family-based linkage study performed on GWAS data suggested a role for rare genetic variants as risk factors for the development of DKD as well [14]. Recently, whole-exome and whole-genome sequencing (WES and WGS, respectively) have enabled the study of low-frequency and rare variants that are expected to have larger effects on the disease risk. WES offers a computationally simpler way of studying protein-altering or truncating variants (PAV and PTV, respectively) [15]. The WGS studies additionally enable exploration of the intronic and intergenic regions, which may affect the gene expression levels through transcription-factor binding-site activity or other regulatory processes [16]. Recently, a WGS in mainly non-diabetic individuals from multiple ancestries identified three novel loci for estimated glomerular filtration rate (eGFR) [17]. However, there are currently only a few WES or WGS-based studies for DKD. Our previous WES study on DKD in T1D yielded no significant findings, whereas a recent WES study identified four exome-wide significant loci for DKD [6, 18]. Furthermore, our previous WGS of 74 sibling pairs proposed involvement of protein kinase C family members with DKD [14, 16]. Here, we study the effect of low-frequency and rare variants on DKD by a WES and WGS meta-analysis design, and perform genome-wide and regulome-wide scans for the non-coding regions using the WGS data (**Figure 1**).

**Figure 1.**
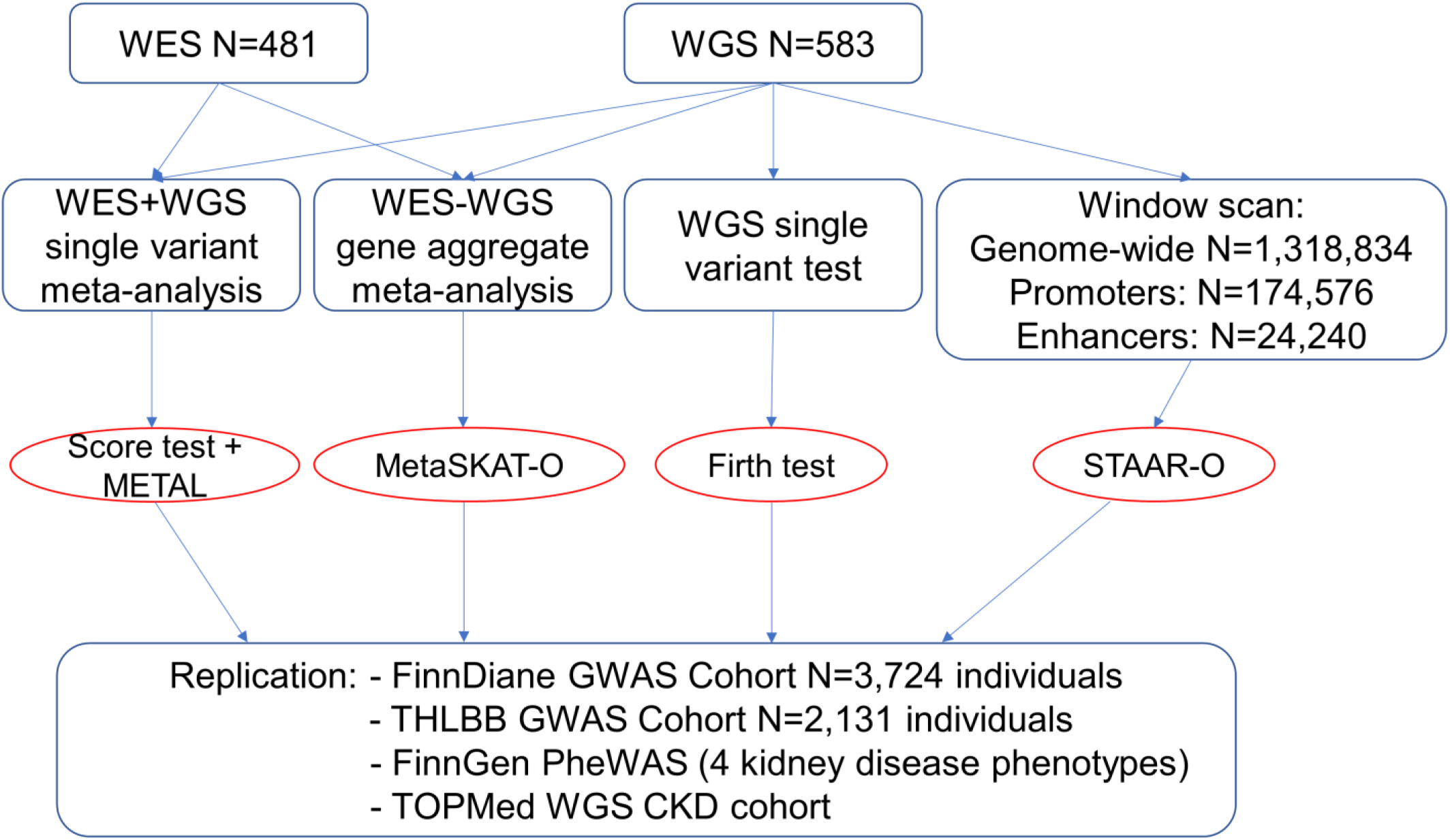
Study setup. The figure illustrates the study setup for single variant and gene- and intergenic region aggregate analyses.

## MATERIALS AND METHODS

### Study population and phenotypes

All participants were recruited from the Finnish Diabetic Nephropathy Study (FinnDiane). Participants were diagnosed with T1D by their attending physician, had an onset of diabetes before the age of 40, and initiated permanent insulin treatment within the first year after the diagnosis (See **Supplementary Methods,** and [19] for more detail). In brief, data on diabetic complications, history of cardiovascular events, and prescribed medications were registered using standardized questionnaires, and blood and urine samples were collected during a standard visit to the attending physician. DKD was defined based on albuminuria status and cases had either severe albuminuria (≥200 μg/min) in two out of three consecutive urine collections, or kidney failure requiring dialysis or a kidney transplant, whereas controls had retained normal albumin excretion rate (AER <20 μg/min) throughout the follow-up.

### Sequencing and data analysis

WES and WGS were performed for 498 and 599 individuals, respectively. WES was performed at the University of Oxford with Illumina HiSeq2000 platform, with an average requirement of 20× target capture with ≥80% coverage as described earlier [6]. WGS was performed with Illumina HiSeq X platform (Macrogen Inc., Rockville, MD, USA), with a requirement of >30× average coverage for mapped reads. Based on the initial quality control (QC) performed at Oxford for WES, or by Macrogen Inc. for WGS, 27 WES samples and 15 WGS samples were excluded due to high homozygosity-heterozygosity ratio, abnormally low mapping depth, or mapped PCR reads.

The WES and WGS samples were processed with the Genome Analysis Toolkit (GATK) 4 Golden Standard pipeline (**Supplementary Methods**) and annotated with SnpEff v5.0e [20, 21]. Post-pipeline variants with <98% call rate and Hardy–Weinberg Equilibrium (HWE) *P*-value <1×10^-10^ were excluded. Principal component analysis with PLINK indicated no population outliers (**Supplementary Figure 1**).

### Variant-based tests

We performed single variant genome-wide association testing for the WGS data with Firth regression, using age of diabetes onset, sex, and the two first genetic principal components as covariates. For the meta-analysis of the WES and WGS data, we performed association testing with inverse variant weighted score test statistic meta-analysis. In WGS single variant analysis, variants with *P*-value <5×10^-8^ were considered genome-wide significant and *P*<5×10^-5^ suggestive. In WES-WGS meta-analysis, *P*<3.5×10^−7^ was considered exome-wide significance and *P*<3.5×10^−5^ as suggestive. All single variant tests were performed with RVTESTS (v2.1.0) and the single variant meta-analysis with METAL v2011-03-25 [22, 23].

### Gene-aggregation tests

We performed gene-based tests with WES and WGS data, separately for protein-truncating variants (PTVs, including frameshift and nonsense variants, and loss or gain of start or stop codons) and protein-altering variants (PAVs, including missense variants, in-frame insertions and deletions (indels), and PTVs), designated to specific genes within RefSeq exomes (**Supplementary Table 1**). Variants were further filtered by their minor allele frequency (MAF) to those with MAF ≤1%, ≤5%, and ≤10%. *P*-values <4×10^−6^ were considered significant (adjusted for 18,226 genes with PAVs with MAF≤10%) and *P*<4×10^−4^ suggestive. Gene-based SKAT- O meta-analysis was performed for WES and WGS cohorts with MetaSKAT v0.8.1 R-package [24].

### Lookup of monogenic kidney disease genes

We studied whether known monogenic kidney disease genes were significantly enriched in our gene-based meta-analysis for DKD. We considered altogether 464 unique genes causing syndromic or monogenic kidney diseases as listed by Connaughton *et al.* [25].

### Whole-genome sliding window and regulome-wide analysis

We performed the genome-wide sliding window tests and aggregation tests for promoters and enhancers for WGS data using STAAR R-package v0.9.6 using the omnibus STAAR-O test [26]. The genome-wide window scan tests were done with window size of 4,000 base pairs (bp) with a gap of 2,000 bps between the windows, and a minimum of 5 variants within the region. As enhancer and promoter regions, we considered the FANTOM5 CAGE profiles (lifted over to GRCh38) (**Supplementary Methods**). At least two variants were required for these regulatory region tests. MAF<5% threshold was used for all the tests. Altogether 174,576 promoters and 24,240 enhancers were tested, resulting in Bonferroni corrected significance levels of *P*<2.86×10^-7^ and 2.12×10^-6^.

Putative enhancer target genes were inspected with Zenbu promoter capture Hi-C data from the YUE Lab database (http://3dgenome.fsm.northwestern.edu/chic.php) [27]. As no Hi-C data were available for kidneys, we searched for bladder as a related tissue.

### Replication

Variants and genes were replicated with FinnDiane GWAS data (**Supplementary Methods**, [28]). Altogether 6,449 individuals and 15.21M variants with imputation quality r^2^> 0.7 passed the QC. After exclusion of individuals included in WES or WGS, the replication was tested in 3,724 individuals with T1D, whereby cases had severe albuminuria or ESRD, and controls had normal AER with minimum of 26 years diabetes duration.

Additionally, replication was attempted in GWAS data for 2,356 non-overlapping Finnish individuals with T1D from the THL Biobank (THLBB) diabetes studies collection (thl.fi/biobank), with registry data for ESRD based on ICD codes available for 2,131 individuals. This GWAS data was imputed with the same pipeline as the main data, using SISu v4 reference panel, and included 16.94M variants after the QC. As albuminuria data were not available for these individuals, they were grouped based on ESRD occurrence — altogether there were 70 individuals with ESRD, and 2,061 individuals without ESRD.

Single variant replication was further tested with Finnish general population GWAS data from the FinnGen project r9 release (https://r9.finngen.fi/) with four kidney disease phenotypes. Finally, we requested replication of the three non-HLA region genes and 16 SNPs in the Trans-Omic for Precision Medicine (TopMED) eGFR WGS study on multi-ancestry general population with 23,732 individuals [17]. P-values <0.0029 were considered Bonferroni significant after correction for altogether 17 tested variants.

### *in silico* annotation of the lead genes and variants

We queried the Human Kidney expression quantitative trait locus (eQTL) Atlas to identify the variants associated with gene expression [29]. Differential gene expression was investigated in kidney conditions data sets in the Nephroseq classic portal v4. Transcription factor Affinity Prediction (TRAP) tool (http://trap.molgen.mpg.de/cgi-bin/trap_two_seq_form.cgi) was used to predict transcription factors with differential binding affinity to the lead regulatory variant reference and alternative allele sequences by searching the JASPAR and TRANSFAC vertebrate motifs with the human background model [30]. Linkage disequilibrium (LD) was evaluated with NIH LDlink tools using the Finnish population (https://ldlink.nih.gov/).

## RESULTS

### Study cohorts

In both the WES and WGS cohorts the cases and controls had similar age of T1D onset and BMI, whereby controls had longer duration of T1D and, by selection, higher baseline eGFR (**Table 1**). All the following analyses were adjusted with sex, age of T1D onset and the two most important genetic principal components.

**Table 1:**
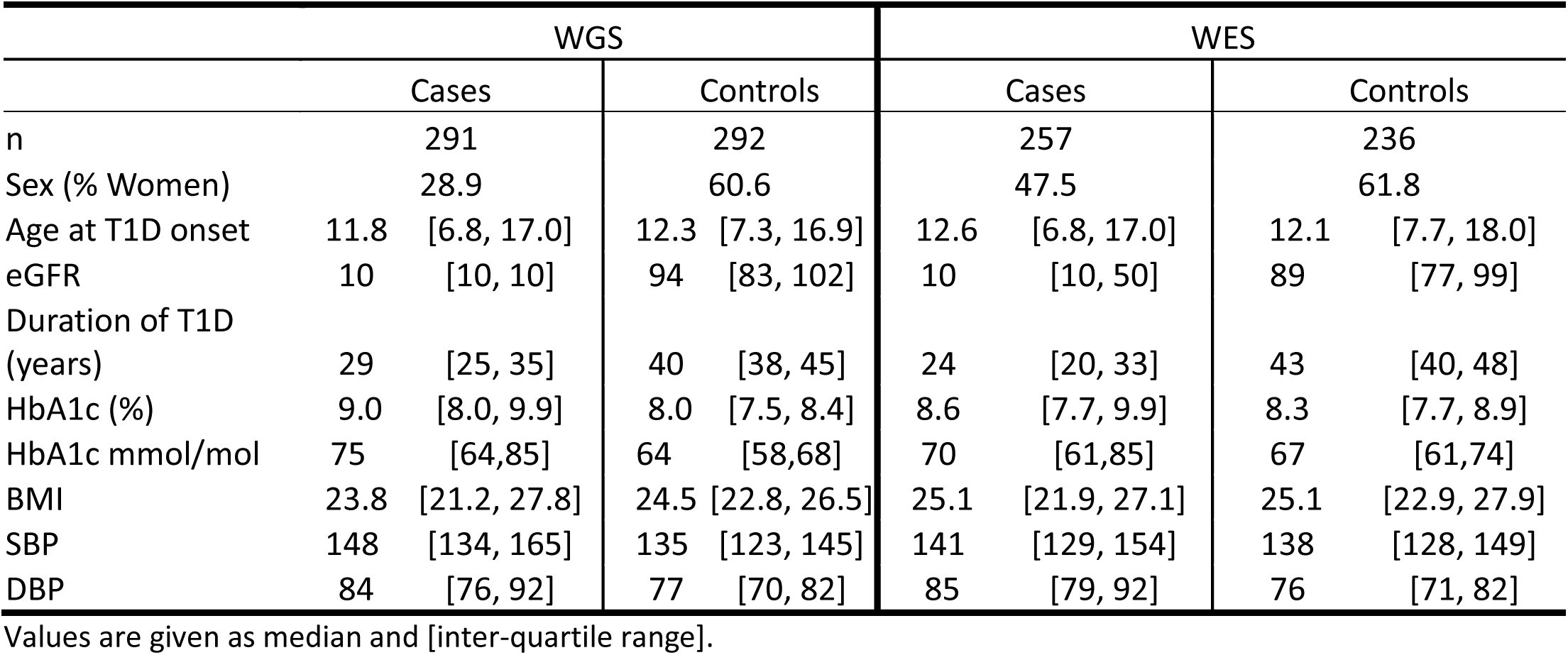
Clinical characteristics for WES and WGS cohorts.

### Single variant associations

The meta-analysis of WES and WGS data resulted in six variants, including two PAVs, with a suggestive *P*-value of 3.5×10^-5^ (**Table 2** and **Figure 2)**. Among these, rs369250 in *THAP7,* and rs1048365 in *AP1S1* showed significant eQTL activity in human kidneys with *THAP7-AS1* (*P*=8.488×10^-45^) and *AP1S1* (*P*=4.98×10^-175^), respectively (**Supplementary Table 2A)**. Extending the analysis outside the exons and their flanking regions, the single variant association test for WGS data identified variants on two intergenic regions 14p12 (close to *CPSF2*) and 16p11.1 (near a group of RNA-genes and pseudogenes), and two intronic variants in *MYO9B* suggestively associated with DKD (*P*-value<5×10^-6^; **Supplementary Table 3**).

**Figure 2.**
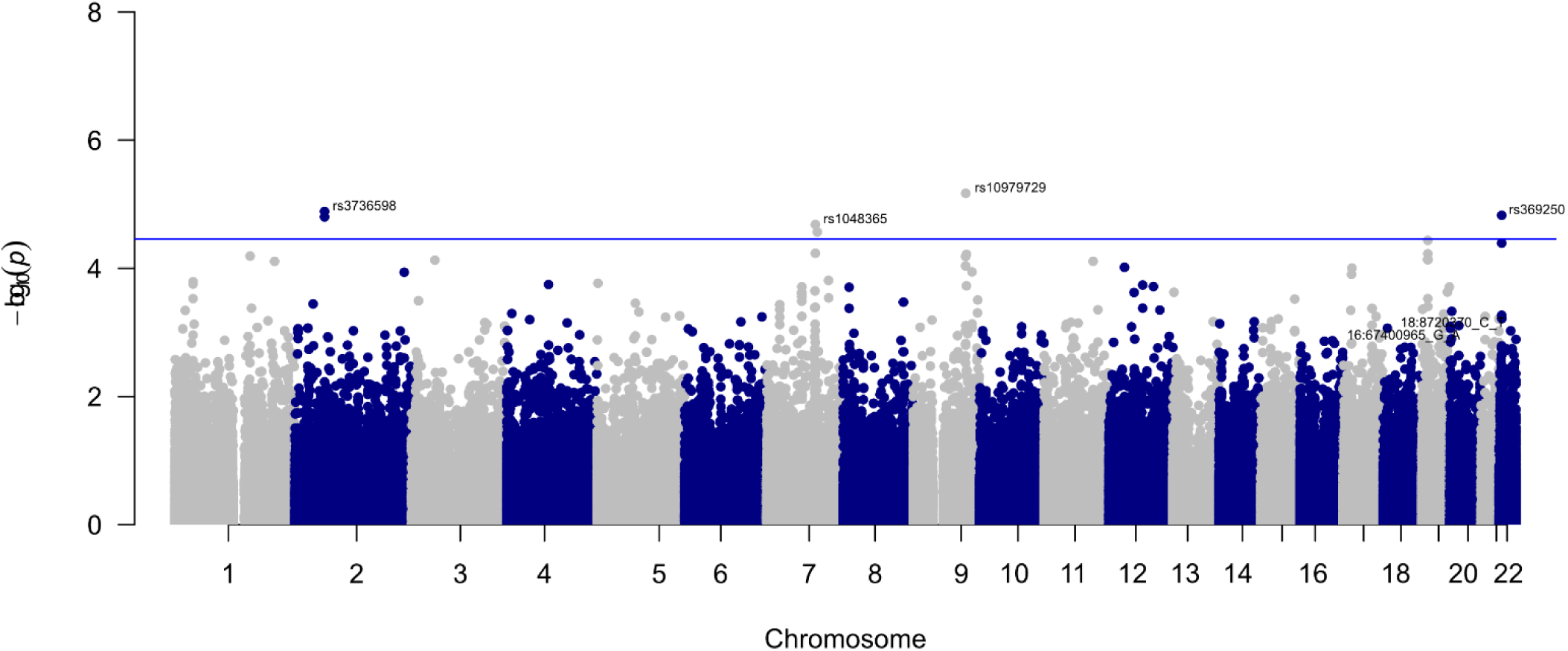
Manhattan plot for DKD WES-WGS single-variant meta-analysis.

**Table 2:** Suggestive associations (p<3.5×10^-5^) in the WES-WGS single variant-based meta-analysis for DKD.

| Position | Variant | Gene | Annotation | MAF | OR | P-value |
| --- | --- | --- | --- | --- | --- | --- |
| 9:109176646 | rs10979729 | *EPB41L4B* | C>T<br>(Pro846Pro) | 0.07 | 0.49 | $6.76 \times 10^{-6}$ |
| 2:61825245 | rs3736598 | *FAM161A* | non-coding | 0.40 | 0.67 | $1.30 \times 10^{-5}$ |
| 22:21002277 | rs369250 | *THAP7* | promoter-TSS | 0.51 | 0.68 | $1.50 \times 10^{-5}$ |
| 2:61826155 | rs6748320 | *FAM161A* | non-coding | 0.40 | 0.67 | $1.58 \times 10^{-5}$ |
| 7:101161149 | rs1048365 | *AP1S1* | 3' UTR | 0.12 | 0.58 | $2.07 \times 10^{-5}$ |
| 7:105107452 | rs117986340 | *KMT2E* | G>T<br>(Gly999Cys) | 0.14 | 1.81 | $2.71 \times 10^{-5}$ |

We attempted replication of the suggestive single variant DKD associations with the non-overlapping FinnDiane and THLBB GWAS data for individuals with T1D (**Supplementary Table 4**), general population FinnGen GWAS data using four kidney disease definitions (**Supplementary Table 5**), and in the TOPMed WGS data for CKD. The *THAP7* promoter rs369250 was replicated for CKD (*P*=2.7×10^-4^, Bonferroni significant) and DKD (*P*=0.012) in the FinnGen GWAS data (**Supplementary Table 5**).

### Gene aggregate tests

In gene-aggregate meta-analysis, *NAT16, LTA, SLC10A6,* and *TSEN54* reached a suggestive *P*-value <4.0×10^−4^ for analyses with low-frequency PAVs (**Table 3**). *NAT16* contained six PAVs with MAF<0.1, and the association was driven by the rs34985488 (A>C p.Phe63Cys, *P*=5.82×10^-5^) (**Supplementary Table 6**). The variant was predicted deleterious or possibly damaging by the SIFT and PolyPhen algorithms, respectively.

**Table 3.** Genes with suggestive *P*<4×10^−4^ in SKAT-O gene aggregate test meta-analysis.

| Gene | P-value | MAF threshold | Variants tested | Variant type | Previous findings |
| --- | --- | --- | --- | --- | --- |
| *NAT16* | $1.4 \times 10^{-4}$ | 0.1 | 6 | PAV+PTV | Significant difference in gene expression between ING patients and controls [31] |
| *LTA* | $1.5 \times 10^{-4}$ | 0.05 | 1 | PAV+PTV | DKD candidate gene [36] |
| *SLC10A6* | $2.7 \times 10^{-4}$ | 0.1 | 3 | PAV+PTV | - |
| *TSEN54* | $3.7 \times 10^{-4}$ | 0.01 | 14 | PAV+PTV | *TSEN2* splice site mutation associated with atypical hemolytic uremic syndrome [41] |

\* *LTA* is localized on HLA-region for which a different variant calling pipeline was used (Please see the Methods for further details).

Replication of the gene aggregate results was sought in FinnDiane GWAS, THLBB GWAS, TOPMed WGS, and UKBB WES gene aggregate data. Furthermore, for the single variants that were nominally significant (*P*<0.05) in the FinnDiane WES+WGS meta-analysis, we tested for replication in the FinnDiane, THLBB, and FinnGen GWAS, and TOPMed WGS data. Of the 4 tested suggestive genes, *NAT16* rs34985488 was replicated in the FinnGen for the CKD phenotype (*P=*0.0028; Bonferroni significant) and *LTA* rs2229092 for the T1D with kidney complications phenotype (*P*=0.0044; rs2229092 was the only identified PAV in *LTA,* ***Supplementary* Table 5**). No replication was observed in the gene aggregate tests (**Supplementary Table 7**).

### Enrichment of Monogenic or syndromic kidney disease genes

We further tested whether monogenic kidney disease genes are enriched among DKD-associated genes (**Supplementary Table 8**). The strongest evidence of enrichment occurred among the “cystic kidney disease or nephronophthisis” class, as 11% (11 out of 96) of the monogenic genes were associated (*P*<0.05) with DKD in our WES-WGS meta-analysis (Binomial test *P* = 0.004).

### Intergenic variant aggregate tests

To improve the statistical power to discover non-coding genetic factors behind DKD, we performed promoter and enhancer aggregate analyses, and functionally-informed genome-wide sliding-window analyses. Two enhancers located at 18q12.3 (*P*=6.78×10^-5^) and 9q21.11 (*P*=2.17×10^-4^) were suggestively associated with DKD (**Table 4**). Based on the Hi-C database, the chromosome 18q12.3 region showed enhancer activity with the closest protein-coding gene *METTL4* (located 186kb upstream) in the bladder. The 9q21.11 locus, close to *CTSL,* is a promoter-enhancer region, which showed enhancer activity with *CTSL* (**Supplementary Table 2C**).

**Table 4.** STAAR-O aggregate analysis results for genome-wide sliding windows (n=1,318,834 regions), promoters (n=184,609) and enhancers (n=24,240)

| WINDOW | VARIANT<br>COUNT | CUMULATIVE<br>MAC | P-VALUE | MAF CLASS | CLOSEST GENE |
| --- | --- | --- | --- | --- | --- |
| SLIDING WINDOWS |  |  |  |  |  |
| chr12:108104001-108108000 (12q14.3) | 14 | 22 | 4.66×10<sup>-6</sup> | 0.01 | WSCD2 |
| chr4:75122001-75126000 4q13.3) | 21 | 61 | 9.46×10<sup>-6</sup> | 0.01 | - |
| PROMOTERS |  |  |  |  |  |
| chr9:87792126-87793126 (9q21.11) | 6 | 79 | 2.67×10<sup>-5</sup> | 0.05 | CTSL3P (Pseudogene) |
| chr2:40755215-40756215 (2q14.2) | 5 | 14 | 3.17×10<sup>-5</sup> | 0.05, 0.01 | LINC01794 (lncRNA) |
| ENHANCERS |  |  |  |  |  |
| chr18:2350865-2351244 (18q12.3) | 2 | 24 | 3.94×10<sup>-5</sup> | 0.05 | METTL4; MYOH1 |
| chr9:87792785-87793201 (9q21.11) | 4 | 27 | 1.17×10<sup>-4</sup> | 0.05 | CTSL |

Cumulative MAC: cumulative minor allele count across all the included variants within the given minor allele frequency (MAF) class.

In the sliding window analysis, the strongest associations were obtained for a chromosomal window on 12q14.3, 21kbps upstream to *WSCD2* (*P*=4.66×10^-6^) and an intergenic region 4q22.3 (*P*=9.46×10^-6^). However, none of the genomic windows (1,318,834 regions), promoter (174,609 regions) or enhancer regions (N=24,240) remained significant after Bonferroni correction (**Table 4**).

Replication for the genome-wide sliding-window, promoter- and enhancer-wide analyses were also performed by testing the nominally significant variants (**Supplementary Table 9**) in the replication data. We observed replication for rs183413211 of the *2q14.2* region 106kb from *SLC8A1* (TOPMed *P*_CKD_ = 0.042; **Supplementary Table 4**), and rs16943099 in the *METTL4* enhancer region in 18q12.3 (FinnGen *P*_T2D_with_renal_compl._=8.6×10^-4^, Bonferroni significant; and *P*_DKD_=0.036; **Supplementary Table 5**).

To assess the potential functional effect of the variants within the identified enhancer regions, we applied the TRAP prediction tool to estimate whether the variants affect the transcription-factor binding probability of the sequence. For the *METTL4*/18q12.3 enhancer region lead SNP, rs1694309, only the reference allele was predicted to provide a binding site for a podocyte-specific transcription factor MafB (TRANSFAC V$MAFB_01, p_REF_ < 1.75×10^-6^, p_ALT_= 0.239; **Supplementary Figure 4**, **Supplementary Table 2D**).

## DISCUSSION

Here, we studied 1,064 Finnish individuals with T1D, representing the extreme phenotypes for DKD, to identify rare and low-frequency variants associated with DKD. We included 546 individuals with severe DKD (severe albuminuria or ESRD) and 528 with T1D and normal albumin excretion rate despite at least 25 years of diabetes duration. We performed single variant meta-analysis for the whole genome including the HLA-region, and we utilized both gene aggregate tests and the regulatory region aggregate test to identify low-frequency variants associated with DKD susceptibility, revealing several putative associations with novel and functionally plausible genes for DKD.

The gene aggregate meta-analysis for low-frequency and rare PAVs and PTVs resulted in four genes suggestively associated with DKD: *NAT16, LTA, SLC10A6,* and *TSEN54*. Through look-ups of the lead SNPs behind the gene aggregate results, replication was found for *NAT16* and *LTA* in the FinnGen cohort (**Supplementary Table 4)**. *NAT16* putatively encodes N-Acetyltransferase 16. N-Acetyltransferases transfer acetyl groups from acetyl-CoA to molecules such as arylamines. According to GTEx RNA-seq data, *NAT16* is expressed in kidney cortex and medulla, along with most other tissues, and *NAT16* was previously observed to show significantly higher expression in individuals with idiopathic nodular glomerulosclerosis compared with healthy controls (Log_2_ fold change=7.11, *P*=1.67×10^-8^) [31]. The lead SNP rs34985488 was classified as deleterious by SIFT and was replicated in the FinnGen CKD phenotype data set (*P=*0.0028). Interestingly, the *NAT16* missense variant rs34985488, and one of the single variant lead SNPs, rs1048365 in the *AP1S1* 3’ untranslated region (UTR), are in moderate LD, and they are both eQTLs for *AP1S1*, *VGF* and *MOGAT3* expression in kidney [29] (**Supplementary Figure 2A**); *MOGAT3* encodes Monoacylglycerol O-acyltransferase 3 that catalyses the synthesis of diacylglycerol from 2-monoacylglycerol and fatty acyl-CoA [32].

The HLA-region gene *LTA* encodes lymphotoxin-alpha (LT-α), also known as tumor necrosis factor β (TNF-β), which plays an important role in the immune response, inflammation, and apoptosis. LT-α binds to TNF receptors, which have been associated with progression of DKD [33, 34]. The *LTA* association with DKD was driven by rs2229092 (OR=0.39, *P*=1.513×10^-5^), and the association was replicated in FinnGen (T1D with kidney complications data set *P*=0.0044). Interestingly, in FinnGen the rs2229092 was protective of T1D (OR=0.74, *P*=3.4×10^-16^), other autoimmune diseases, and diabetic complications, ophthalmic complications being the most significant OR=0.70, *P*=4.5×10^-13^ (**Supplementary Table 4**). However, others have noted that assessing causal effects of rs2229092 is difficult due to pleiotropy [35]. Of note, an association between DKD and *LTA* p.T60N (rs1041981) was suggested by candidate gene studies already 15 years ago, but the association has not been replicated in more recent large GWASs [36]. This could be partly due to leaving out the HLA-region from GWAS imputation or sequencing due to complexity of the region.

The WGS data allowed us to investigate low-frequency variants also in the non-coding regions. On chromosome 18q12.3, enhancer interacting with *METTL4* gene was suggestively associated with DKD (*P*=6.78×10^-5^). The association was led by rs16943099, and it replicated for “T2D with kidney complications” (*P=8.6*×10^-4^) in the FinnGen GWAS data. *In silico* prediction suggested that the rs16943099 minor C allele, associated with lower risk of DKD, disrupted a transcription-factor binding site for a podocyte-specific transcription factor Mafb (**Supplementary Figure 3**, **Supplementary Table 2D**); forced *mafb* expression was recently shown to prevent CKD in mice [37]. *METTL4* encodes Mettl4 methyltransferase, which has been shown to mediate m^6^Am methylation on U2 snRNA *in vitro* [38]. An intergenic variant rs185299109 in the *LINC00470/METTL4* locus was previously associated with DKD (eGFR based CKD phenotype, *P*=1.3×10^-8^) [10]. Moreover, methyl adenosine modification of paralogous *METTL3* was observed to promote podocyte injury in DKD [39], and a rare intronic variant in *METTL8* was one of the novel findings for eGFR in the TOPMed WGS [17].

The 9q21.11 enhancer (*P*=1.17×10^-4^) and the partly overlapping promoter (*P*=2.67×10^-5^) are adjacent to *CTSL,* encoding Cathepsin L. Cathepsins are proteases which degrade and detoxify advanced glycation end product (AGE)-modified proteins. Higher concentrations of urinary cathepsin L were predictive of improvement in albuminuria after four years of follow-up, and positively correlated with urinary glucosepane, a cross-linking AGE derived from D-glucose, in individuals with type 2 diabetes [40].

To date, our WES/WGS of 1,097 individuals with T1D is one of the largest sequencing studies for DKD. However, previous studies have shown that a larger sample size is often needed to discover variants with modest effect size, and thus, the main limitation of this study is the sample size. However, the study participants were carefully selected and characterized for their phenotype, and had either advanced DKD or long duration of T1D without DKD. We could not replicate common variants previously identified for DKD, such as the reported *COL4A3* missense variant rs55703767 (*P*=0.056), suggesting that we may have missed variants relevant for DKD due to the limited sample size. However, our focus here was on discovery of rare variants with functional relevance.

We used two different sequencing platforms in the study. Even though the datasets were analysed using the same pipeline, there were differing read lengths of 150 bps for WGS, and 100 bps for WES, and the mean amount of low-frequency variants per gene was significantly greater in the WGS (5.88) compared to the WES data set (5.31). Due to the limitation of databases and tools, our study included only transcribed enhancers, and the promoters were defined with an arbitrarily selected 1,000 bp extension downstream TSS, although the promoter lengths vary.

Finally, no other WGS data for DKD were available for replication of our findings, but we attempted replication in multiple data sets, including studies with imputed GWAS data for DKD in T1D, and GWAS, WES, and WGS data sets of phenotypes gathered from the general population for kidney disease. Due to these limitations, we here report replication at nominal significance (*P*<0.05). After correcting for the total number of tested SNPs (n=17), 18q12.3 (*METTL4*) rs16943099, *THAP7* rs369250, and *NAT16* rs34985488 remained significant. In addition, evidence from eQTL and differential gene expression in kidney tissue supports the relevance of the novel loci identified here, especially for the 18q12.3 (*METTL4*) enhancer region, and *NAT16* and *LTA* genes.

## CONCLUSIONS

In conclusion, this WES-WGS meta-analysis resulted in several plausible DKD genes that may be important for the development of DKD in individuals with T1D. However, further validation is needed to confirm their role in DKD.

## Supporting information

The supplements should be placed after the main manuscript

## Acknowledgements

We are indebted to the late Carol Forsblom (1964–2022), the international coordinator of the FinnDiane Study Group, for his considerable contribution. The skilled technical assistance of H. Krigsman, H Olanne, M. Parkkonen, M. Korolainen, A. Sandelin, J. Tuomikangas, and Kirsi Uljala (Folkhälsan Research Center, Finland) are gratefully acknowledged. The authors also acknowledge all the physicians and nurses at each center taking part in the enrolment and clinical characterization of the participants (see **Supplementary table 10** for a list of study centres and investigators involved in the FinnDiane study). The THLBB GWAS data used for the research were obtained from the THL Biobank (study number THLBB2021_10). We thank all study participants for their generous participation in the THL Biobank and THL Diabetes Studies. We acknowledge that the ELIXIR Finland node, hosted at the CSC – IT Center for Science for ICT resources, enabled the WES and WGS data processing.

## Funding

This study was supported by funding from Folkhälsan Research Foundation, Wilhelm and Else Stockmann Foundation, Liv och Hälsa Society, Munuaissäätiö, Helsinki University Hospital Research Funds (EVO TYH2018207), Academy of Finland (299200, and 316664), Novo Nordisk Foundation (NNF OC0013659, NNF23OC0082732), the Sigrid Jusélius Foundation, and the Finnish Diabetes Research Foundation.

## Author contributions

JKH performed data processing and statistical analysis, and wrote the original draft. AAA performed data processing and did intergenic variant analysis pipeline. NS contributed to data analysis and investigation, edited the manuscript, and supervised the work. AS contributed to GWAS data processing and edited the manuscript. EV contributed to WGS patient selection and reviewed the manuscript, ED performed the THLBB data processing, P-HG supervised the work, BLM and NF performed the replication in TOPMed study. All authors approve the version to be submitted published, and agree to be accountable for all aspects of the work in ensuring that questions related to the accuracy or integrity of any part of the work are appropriately investigated and resolved.

## Data availability statement

The FinnDiane WES and WGS datasets generated and/or analyzed during the current study are not publicly available as the participants’ written consent does not allow data sharing. The Readers may propose collaboration to research the individual level data with correspondence with the lead investigator.

## Disclosures

P-HG has served on advisory boards for AbbVie, Astellas, AstraZeneca, Bayer, Boehringer Ingelheim, Eli Lilly, Janssen, Medscape, MSD, Mundipharma, Novartis, Novo Nordisk, Sanofi, and has received lecture honoraria from Astellas, AstraZeneca, Bayer, Boehringer Ingelheim, Eli Lilly, Elo Water, Medscape, MSD, Mundipharma, Novartis, Novo Nordisk and Sanofi. P-H G has also received investigator-initiated grants from Eli Lilly and Roche.

## Notes

### Funding Statement

This study was supported by funding from Folkhalsan Research Foundation, Wilhelm and Else Stockmann Foundation, Liv och Halsa Society, Munuaissatio, Helsinki University Hospital Research Funds (EVO TYH2018207), Academy of Finland (299200, and 316664), Novo Nordisk Foundation (NNF OC0013659, NNF23OC0082732), the Sigrid Juselius Foundation, and the Finnish Diabetes Research Foundation.

### Author Declarations

This study was performed in accordance with the Declaration of Helsinki. Ethics committee of the Helsinki and Uusimaa Hospital District (491/E5/2006, 238/13/03/00/2015, and HUS-3313-2018) have given ethical approval for the study protocol, and all participants gave informed consent before participation.

