## Supplementary material for "Whole-exome and Whole-genome Sequencing of 1097 Individuals with Type 1 Diabetes Reveals Novel Genes for Diabetic Kidney Disease": The supplements should be placed after the main manuscript

ELECTRONIC SUPPLEMENTARY MATERIAL

**Supplementary Figures: 4**

**Supplementary Tables: 10**

**Supplementary Methods**

### Patients

The Finnish Diabetic Nephropathy Study (FinnDiane) is an ongoing nationwide prospective study, established in 1997 to pinpoint risk factors for long-term diabetic complications. The study currently comprises more than 6000 individuals with T1D. The study protocol was approved by the Ethical Committee of the Helsinki and Uusimaa Hospital District (491/E5/2006, 238/13/03/00/2015, and HUS-3313-2018, July 3rd 2019) and participants gave their informed consent before recruitment. This study was performed following the Declaration of Helsinki.

### Sequencing data analysis with GATK pipeline

First, the fastq-reads were trimmed with Trimmomatic v0.36, and the trimmed reads were run through FastQC v0.11.9 and results were aggregated and assessed with MultiQC v1.11. The reads were aligned by lanes (1-8 lanes in WGS and 1-2 lanes in WES), sorted, and removed duplicates with Picard's SortSam and MarkDuplicates tools. The reads were recalibrated by chromosome with GATK BQSR and ApplyBQSR tools, and variants were called with HaplotypeCaller tool's ERC mode into a single sample GVCF-file. The GVCF-files were combined into a multi-sample GVCF with GATK CombineGVCFs tool and transformed into a VCF-file with GenotypeGVCFs tool, separately for WES and WGS samples. Variants were then filtered using excess heterozygosity threshold of 54.69. SNPs and indels were filtered separately according to tranche thresholds recommended by GATK, and truth sensitivity level of 99.7%. All variants were annotated with SnpEff v5.0e based on GRCh38.99 database. Comparison with pre-existing GWAS genotyping showed 99.5% concordance with the sequencing data. To prepare chromosome with ALT-contigs (HLA-region) we used a workflow from https://gatk.broadinstitute.org/hc/en-us/articles/360037498992--How-to-Map-reads-to-a-reference-with-alternate-contigs-like-GRCH38 with modifications for GATK4.

**Replication of findings with FinnDiane GWAS**

Genomes were genotyped in four batches at the University of Virginia with HumanCoreExomBead arrays 12-1.0, 12-1.1, and 24-1.0 (Illumina, San Diego, CA, USA). The GWAS data were lifted over from GRCh37 to GRCh38 genetic coordinates using previously described pipeline, and sample data from the four genotyping batches were merged [1]. We removed individuals with high genotype missingness (>5%) and variants with excess heterozygosity (±4 standard deviations), high missingness (>2%), low HWE *P*-value ($<10^{-6}$), or minor allele count <3. For imputation, chip-genotyped samples were pre-phased with Eagle 2.3.5, and genotype imputation was performed with Beagle 4.1 (version 08Jun17.d8b) based on the population-specific SISu v3 reference panel, consisting of WGS data for 3,775 Finnish individuals. Variants were annotated with snpEff5.0e [2]. Single variant replications were conducted using Firth test with RVTESTS and gene aggregate replications SKAT-O test with REGENIE [3].

**Whole-genome sliding window analysis and regulome-wide analysis**

Window length was extended from default (2000 bp) to increase statistical power. Importantly, sliding window analyses were conducted by weighting the variants by Combined Annotation Dependent Depletion (CADD) v1.6 functional annotations, variant rarity, and annotation principal components (a-PC), which were calculated previously, following Li et al. (2020) guideline [4, 5]. The CAGE transcription stat sites (TSSs) were extended to form full-length promoters (1000 bp), while variants within enhancer and promoters were weighted only according to variant rarity – a functional annotation available for all variants.

**Supplementary Figure 1 - Genetic principal components (PC) 1 and 2 plotted for A WGS and B WES** suggest that after the sample and variant QC, there were no significant outlier samples and no detectable ancestry clusters. The PC1 explained 0.69% and PC2 0.63% of variance within WES and WGS.

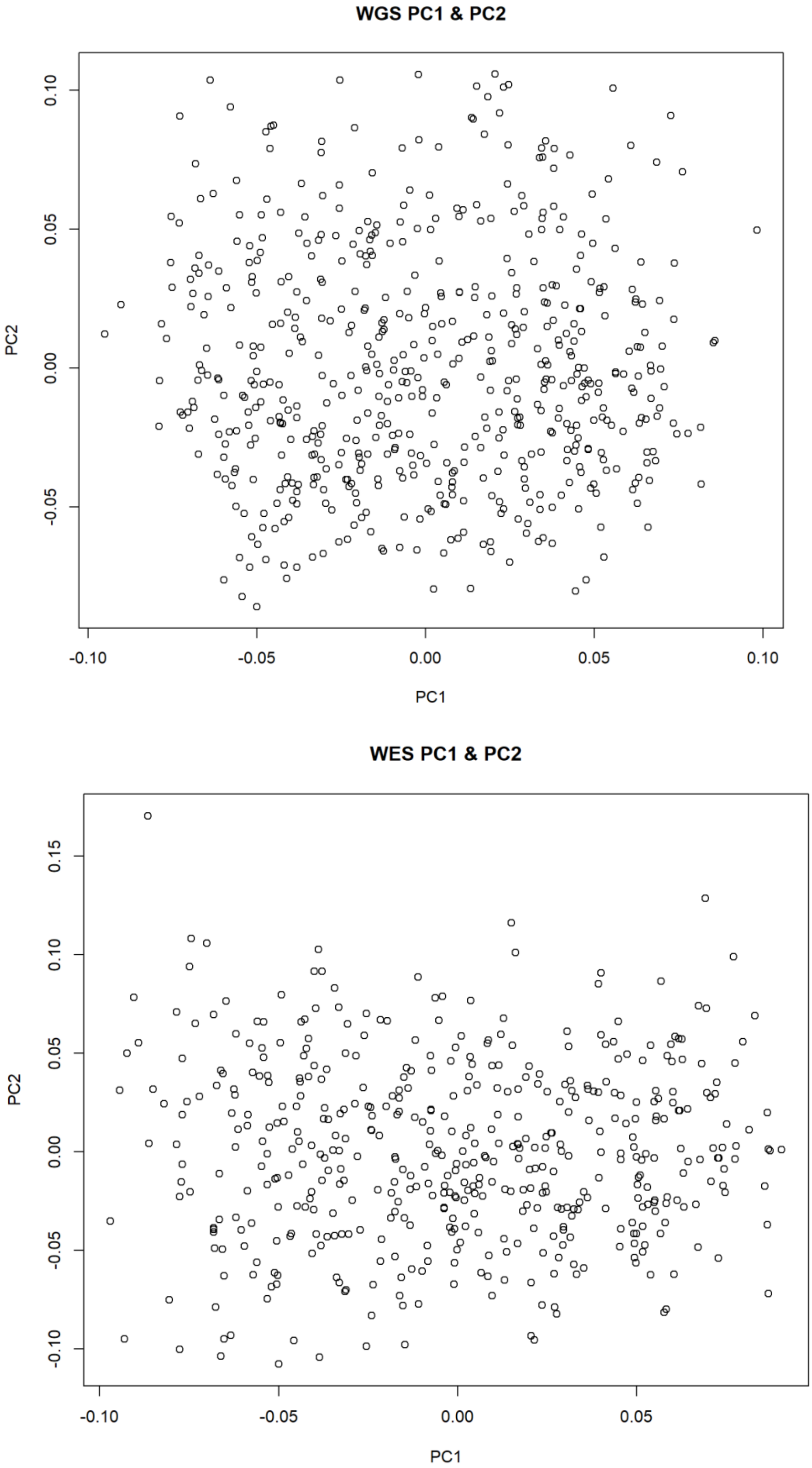

**Supplementary Figure 2 – Locuszoom on chromosome 7:101.0-101.5Mb**. The highlighted *AP1S1* 3' UTR rs1048365 and *NAT16* p.Phe63Ser rs34985488 both have significant eQTL activity in tubule on AP1S1, VGF and MOGAT3 (colored in orange).

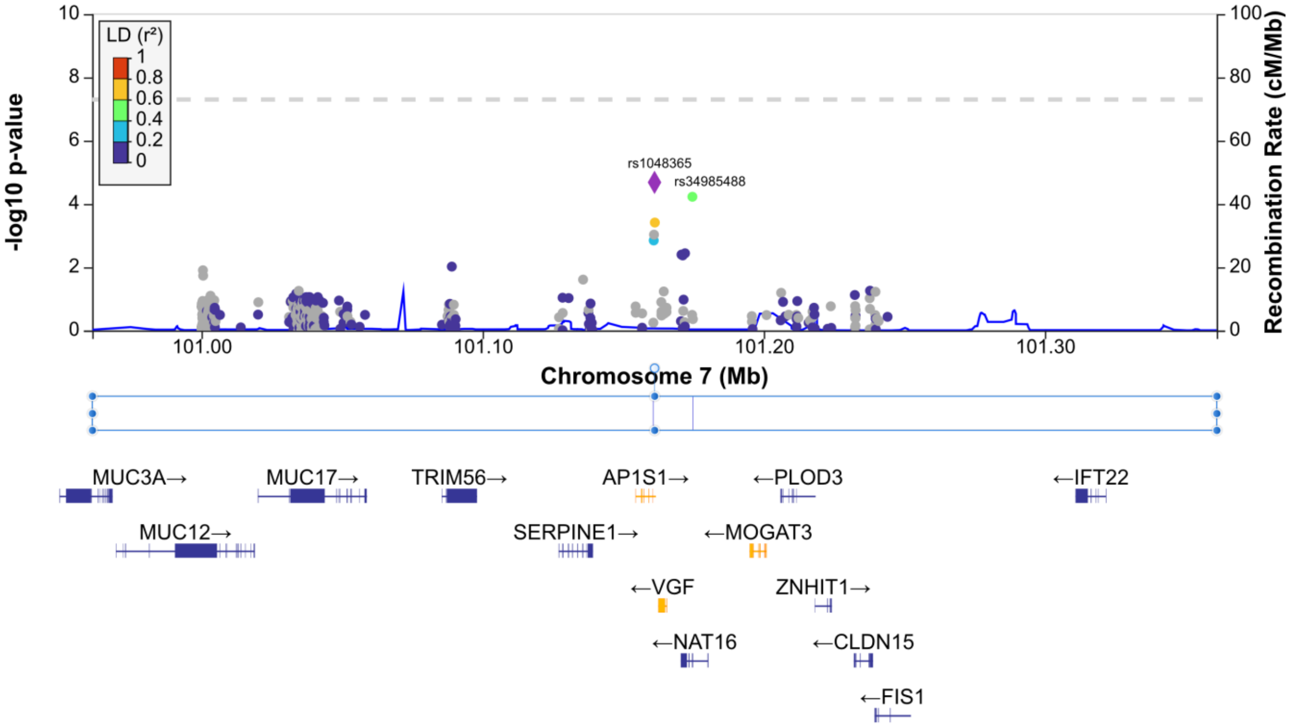

**Supplementary Figure 3 – METTL4 enhancer region rs1694309* is associated with DKD and alters Mafb binding affinitiy** (**A**). Red color indicates TRAP predicted binding region for the JASPAR MA0117.1 motif. **B**: MAFB and METTL4 gene expression in human kidneys (Wilkinson et al., 2019). Figures plotted at [http://humphreyslab.com/SingleCell/](http://humphreyslab.com/SingleCell/). **C** and **D**: in nephroseq database, Woroniecka et al. data for DKD vs healthy living donors, *MAFB* is underexpressed in DKD in glomeruli (**C**; p=4.6×10-4, fold change -4.17) and overexpressed and tubuli (**D**; p=4.9×10-4, fold change 2.25).

**A:**

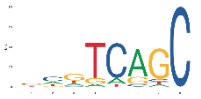

REF tattcattcaaaacaggattgagACGTCAGCttgacaaagctttctctcct
ALT tattcattcaaaacaggattgagacctcagcttgacaaagctttctctcct
*

**B:**

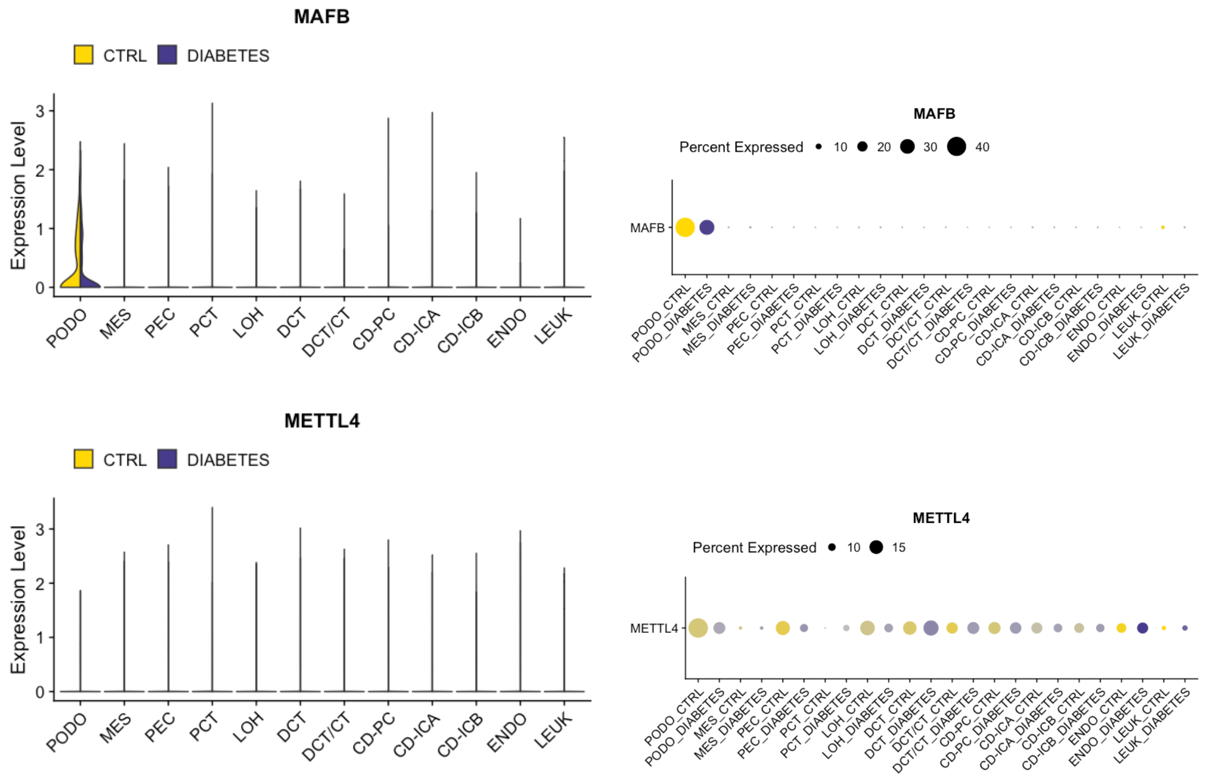

**C:** **D:**

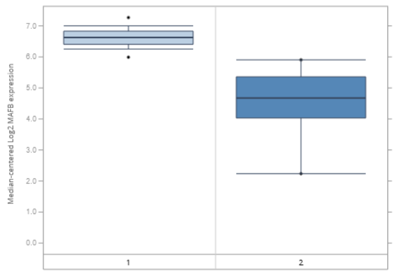

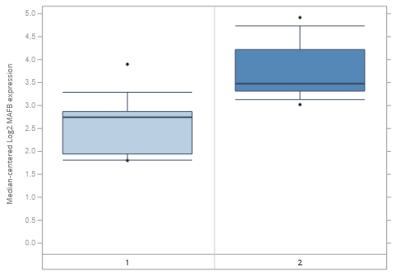

**Supplementary Table 1 – Variant function classes from snPEff annotation, used for the gene aggregate test**. Protein-truncating variants (PTVs) include variants that are expected to severely or completely disrupt protein function, whereas protein-altering variants include the PTV, but also variants, such as missense variants and INDELs, that are expected to have less effect on protein function

| **Protein-altering variants (PAVs)** | **Protein-truncating variants (PTVs)** |
|---|---|
| 5_prime_UTR_premature_start_codon_gain_variant | start_lost |
| 5_prime_UTR_truncation&exon_loss_variant | stop_gained |
| bidirectional_gene_fusion | stop_lost |
| gene_fusion | bidirectional_gene_fusion |
| conservative_inframe_deletion | gene_fusion |
| conservative_inframe_insertion | frameshift_variant |
| disruptive_inframe_deletion | exon_loss_variant |
| disruptive_inframe_insertion | splice_acceptor_variant |
| start_lost | splice_donor_variant |
| stop_gained | |
| stop_lost | |
| exon_loss_variant | |
| frameshift_variant | |
| missense_variant | |
| splice_acceptor_variant | |
| splice_donor_variant | |
| structural_interaction_variant | |

**Supplementary Table 2 – eQTL, gene expression and HI-C capture data obtained from databases. A** Human kidney eQTL data for the top single variant analysis results. Top WES-WGS meta-analysis genes and genome-wide sliding windows, promoters and enhancers (for which variants with $P<0.05$ were tested) from the Human Kidney eQTL Atlas, **B** gene expression for the top meta-analysis genes on kidney other kidney conditions obtained from Nephroseq database, **C** Hi-C capture the top enhancer regions (SNPs with p<0.05 within regions were queried) **D** Predicted allele-specific Mafb binding activity of rs1694309, when the ±25bp sequence was queried with Transcription factor Affinity Prediction (TRAP) tool and jasper vertebrates and transfac_2010.1 vertebrates (human motifs only) databases.

**A**

| SNP | Position | Gene | Direction | $Effect_{ALT}$ | SE | P |
|---|---|---|---|---|---|---|
| rs1048365 | 7:100804430:C_T | *AP1S1* | ++++ | 0.9991 | 0.0354 | $4.98\times10^{-175}$ |
| rs1048365 | 7:100804430:C_T | *VGF* | ++++ | 0.6245 | 0.0552 | $1.147\times10^{-29}$ |
| rs1048365 | 7:100804430:C_T | *MOGAT3* | ++-+ | 0.2295 | 0.0473 | $1.235\times10^{-6}$ |
| rs34985488 | 7:100817901:A_G | *AP1S1* | +?++ | 0.9021 | 0.0584 | $8.056\times10^{-54}$ |
| rs34985488 | 7:100817901:A_G | *VGF* | +?++ | 0.441 | 0.0741 | $2.646\times10^{-9}$ |
| rs34985488 | 7:100817901:A_G | *MOGAT3* | +?-+ | 0.3036 | 0.058 | $1.644\times10^{-7}$ |
| rs369250 | 22:21356566:A_G | *THAP7-AS1* | -?-- | -0.6864 | 0.0489 | $8.488\times10^{-45}$ |

**B**

| PHENOTYPE | GENE | N (CASES/CONTROLS) | P-VALUE | T-TEST | FOLD-CHANGE | NOTES |
|---|---|---|---|---|---|---|
| NAKAGAWA CKD KIDNEY | *MTHFR* | 48/5 | $1.03\times10^{-18}$ | -14.684 | -4.773 | |
| MEMBRANOUS GLOMERULONEPHROPATHY | *LTA* | 9/3 | 0.002 | 5.589 | -1.641 | |
| HODGIN DIABETES MOUSE GLOM | *SLC10A6* | 7/5 | 0.002 | 3.894 | 2.334 | |
| NAKAGAWA CKD KIDNEY | *TSEN54* | 48/5 | $1.90\times10^{-14}$ | -10.582 | -5.181 | Controls have increased expression of TSEN54, about half of cases have decreased, half increased expression |

**C**

| SNP | Tissue | Interacting regions (hg19) |
|---|---|---|
| rs16943099 | Bladder | chr18:2349726-2356087 (enhancer): chr18:2567644-2580208 (*METTL4*) |
| rs34407883 | Bladder | chr9:90395315-90407069 (enhancer) : chr9:90335670-90343626 (*CTSL*) |

**D**

| **Database for Transcription factor binding site motifs** | **$p_{REF}$** | **$p_{ALT}$** | **Rank** | **Difference log(p) for two sequences** | **Matrix ID** | **Matrix name** |
|---|---|---|---|---|---|---|
| **rs16943099 (G/C) (18q12.3)** | | | | | | |
| TRANSFAC vertebrates | $<1.75\times10^{-6}$ | 0.239 | 1 | 5.13 | V$MAFB_01 | Mafb |
| jaspar vertebrates | $6.3\times10^{-4}$ | 0.255 | 1 | 2.61 | MA0117.1 | Mafb |
| **rs12349926 (G/A) (9q21.11)** | | | | | | |
| TRANSFAC vertebrates | 0.004 | $<4\times10^{-6}$ | 1 | -3.03 | M01665 | Irf8_Q6 |
| jaspar vertebrates | 0.61 | 0.16 | 1 | -0.59 | MA0028.1 | ELK1 |

**Supplementary Table 3 – WGS single variant analysis for DKD using Firth test**

| Position | rs# | Genes* | OR | AF | P-value | Replication P-value (FD GWAS) |
|---|---|---|---|---|---|---|
| 14:92202604 | rs66867671 | Close to *CPSF2* | 0.50 | 0.32 | $1.212\times10^{-6}$ | 0.846 |
| 14:92203776 | rs12881014 | Close to *CPSF2* | 0.51 | 0.32 | $1.658\times10^{-6}$ | 0.814 |
| 14:92206163 | rs61976635 | Close to *CPSF2* | 0.51 | 0.32 | $1.898\times10^{-6}$ | 0.814 |
| 14:92208383 | rs35205959 | Close to *CPSF2* | 0.52 | 0.33 | $4.016\times10^{-6}$ | 0.880 |
| 14:92208614 | rs61976640 | Close to *CPSF2* | 0.52 | 0.33 | $4.016\times10^{-6}$ | 0.887 |
| 14:92209896 | rs11624199 | Close to *CPSF2* | 0.53 | 0.32 | $6.237\times10^{-6}$ | 0.901 |
| 16:36109142 | . | Multiple RNA/ pseudogenes, closest to *AC116553.3* | 2.84 | 0.10 | $4.362\times10^{-6}$ | - |
| 16:36109143 | . | Multiple RNA/ pseudogenes, closest to *AC116553.3* | 2.74 | 0.10 | $8.422\times10^{-6}$ | - |
| 19:17174120 | rs181653048 | *MYO9B* intron | 0.39 | 0.11 | $9.395\times10^{-6}$ | 0.794 |
| 19:17174122 | rs112921283 | *MYO9B* intron | 0.39 | 0.11 | $9.395\times10^{-6}$ | 0.794 |

**Supplementary Table 4 – Single variant analysis replication in FinnDiane and THL GWASes for DKD, TOPMed WGS for CKD**

| POSITION | RSID | GENE* | CADD | SIFT | POLYPHEN | TOPMED MAF | TOPMED P | FINNDIANE GWAS P | THLBB GWAS P |
|---|---|---|---|---|---|---|---|---|---|
| **SINGLE VARIANT META-ANALYSIS** | | | | | | | | | |
| 2:61825245 | rs3736598 | *FAM161A* | 11.7 | - | - | 0.38 | 0.19 | 0.43 | 0.68 |
| 2:61826155 | rs6748320 | *FAM161A* | 1.8 | - | - | 0.38 | 0.19 | 0.43 | 0.68 |
| 22:21002277 | rs369250 | *THAP7* | 5.27 | - | - | 0.44 | 0.71 | 0.17 | 0.61 |
| 7:101161149 | rs1048365 | *AP1S1* | 2.75 | - | - | 0.25 | 0.92 | 0.76 | NA |
| 7:105107452 | rs117986340 | *KMT2E* | 27.5 | deleterious_lc | probably_damaging | 0.03 | 0.87 | 0.60 | 0.97 |
| 9:109176646 | rs10979729 | *EPB41L4B* | 1.43 | - | - | 0.05 | 0.12 | 0.52 | 0.61 |
| **GENE AGGREGATE META-ANALYSIS SNPS** | | | | | | | | | |
| 17:75522195 | rs200434678 | *TSEN54* | 24.9 | deleterious | probably_damaging | 0.0003 | 0.62 | 0.89 | NA |
| 17:75523335 | rs200228117 | *TSEN54* | 25.4 | Tolerated | benign | NA | NA | 0.29 | 0.69 |
| 4:86849099 | rs17694522 | *SLC10A6* | 22.5 | deleterious | benign | 0.04 | 0.08 | 0.79 | 0.86 |
| 6:31572980 | rs2229092 | *LTA* | 6.52 | tolerated | benign | 0.04 | 0.52 | 0.27 | 0.35 |
| 7:101174620 | rs34985488 | *NAT16* | 25.4 | deleterious | possibly_damaging | 0.15 | 0.75 | 0.58 | 1.00 |
| **SLIDING-WINDOW, PROMOTOR AND ENHANCER-WIDE ANALYSIS ON WGS DATA** | | | | | | | | | |
| 18:2351193 | rs16943099 | *METTL4* | 0.651 | - | - | 0.01 | 0.38 | 0.79 | 0.72 |
| 2:40755782 | rs183413211 | *LINC01794* | 2.10 | - | - | 0.004 | **0.04** | 0.66 | 0.64 |
| 4:75124427 | rs114761270 | - | 9.12 | - | - | 0.007 | 0.50 | 0.46 | 0.25 |
| 9:87792152 | rs34407883 | *CTSL* | 0.837 | - | - | 0.001 | 0.98 | NA | NA |
| 9:87792755 | rs12349827 | *CTSL* | 0.305 | - | - | 0.01 | 0.25 | NA | 0.94 |
| 9:87793063 | rs12349926 | *CTSL* | 0.538 | - | - | 0.03 | 0.89 | 0.37 | 0.71 |

* Underlying, annotated or closest gene

**Supplementary Table 5 – Variant replication across FinnGen kidney disease phenotypes**

| POSITION | RSID | GENES/ REGION | CKD P | DIABETIC NEPHROPATHY P | T1D WITH RENAL COMPLICATIONS P | T2D WITH REAL COMPLICATIONS | OTHER |
|---|---|---|---|---|---|---|---|
| **SINGLE VARIANT META-ANALYSIS** | | | | | | | |
| 9:109176646 | rs10979729 | *EPB41L4B* | 0.88 | 0.53 | 0.085 | 1.0 | |
| 2:61825245 | rs3736598 | *FAM161A* | 0.46 | 0.26 | 0.96 | 0.33 | |
| 22:21002277 | rs369250 | *THAP7* | **$2.7\times10^{-4}$** | **0.012** | 0.27 | **0.01** | |
| 2:61826155 | rs6748320 | *FAM161A* | 0.47 | 0.26 | 0.98 | 0.33 | |
| 7:101161149 | rs1048365 | *AP1S1* | NA | NA | NA | NA | |
| 7:105107452 | rs117986340 | *KMT2E* | 0.81 | 0.93 | 0.44 | 0.68 | |
| **GENE AGGREGATE META-ANALYSIS SNPS** | | | | | | | |
| 4:86849099 | rs17694522 | *SLC10A6* | 0.97 | 0.27 | 0.71 | 0.35 | |
| 6:31572980 | rs2229092 | *LTA* | 0.21 | **0.027** | **0.0044** | 0.88 | T1D, wide definition P=3.4e-16, T1D with ophthalmic complications P=4.5e-13 |
| 7:101174620 | rs34985488 | *NAT16* | **0.0028** | 0.16 | 0.32 | 0.48 | |
| 17:75522195 | rs200434678 | *TSEN54* | 0.58 | 0.87 | 0.60 | 0.96 | |
| 17:75523335 | rs200228117 | *TSEN54* | 0.65 | 0.75 | 0.97 | 0.71 | |
| **SLIDING-WINDOW, PROMOTOR AND ENHANCER-WIDE ANALYSIS ON WGS DATA** | | | | | | | |
| 9:87793063 | rs12349926 | *CTSL* | 0.63 | 0.13 | 0.26 | 0.22 | |
| 9:87792152 | rs34407883 | *CTSL* | NA | NA | NA | NA | |
| 9:87792755 | rs12349827 | *CTSL* | 0.98 | 0.17 | 0.56 | 0.24 | |
| 2:40755782 | rs183413211 | *LINC01794* | 0.27 | 0.72 | 0.96 | 0.30 | |
| 4:75124427 | rs114761270 | - | 0.27 | 0.28 | 0.37 | 0.67 | |
| 18:2351193 | rs16943099 | *METTL4* | 0.10 | **0.036** | 0.17 | **$8.6\times10^{-4}$** | |

**Supplementary Table 6 – Individual associations of low frequency PAVs and PTVs inside suggestively DKD-associated genes**

| GENE (VARIANT FILTERS) | RSID | POSITION | ALLELE1 | ALLELE2 | MAF WGS | MAF WES | ZSCORE | P-VALUE | DIRECTION WES/WGS) |
|---|---|---|---|---|---|---|---|---|---|
| ***NAT16*** **(PAV 10%)** | | 7:101172173 | a | g | MAC≤3 | | 1.399 | 0.1617 | ?+ |
| | rs781324827 | 7:101172525 | a | c | MAC≤3 | | 0.661 | 0.5086 | ?+ |
| | rs768356640 | 7:101172554 | g | ggcagcggcgagaa ggtgccagag gtcc | MAC≤3 | | -1.327 | 0.1845 | ?- |
| | | 7:101174511 | a | c | | MAC≤3 | -0.995 | 0.3197 | -? |
| | rs34985488 | 7:101174620 | a | g | 0.09862 | 0.07449 | 4.016 | **5.916e-05** | ++ |
| | | 7:101174795 | a | c | | MAC≤3 | -0.782 | 0.4343 | -? |
| ***SLC10A6*** **(PAV 10%)** | rs139158966 | 4:86823730 | c | g | MAC≤3 | MAC≤3 | -1.116 | 0.2646 | -- |
| | rs770266438 | 4:86831834 | a | c | MAC≤3 | | 0.715 | 0.4745 | ?+ |
| | rs17694522 | 4:86849099 | a | g | 0.06861 | 0.07347 | 3.714 | **0.000204** | ++ |
| ***TSEN54*** **(PAV 1%)** | | 17:75517212 | a | c | MAC≤3 | | -0.735 | 0.4623 | ?- |
| | rs758995350 | 17:75517582 | a | c | | MAC≤3 | 1.153 | 0.249 | +? |
| | rs770897552 | 17:75519014 | c | g | MAC≤3 | | 0.638 | 0.5232 | ?+ |
| | rs200015685 | 17:75521847 | a | g | MAC≤3 | | 1.358 | 0.1746 | ?+ |
| | rs762725861 | 17:75521901 | a | g | MAC≤3 | | -0.735 | 0.4625 | ?- |
| | rs777546135 | 17:75521908 | t | g | | MAC≤3 | -0.883 | 0.3772 | -? |
| | | 17:75521978 | t | g | | MAC≤3 | -1.02 | 0.3075 | -? |
| | | 17:75522000 | t | g | | | -1.199 | 0.2305 | -? |
| | | 17:75522059 | t | c | MAC≤3 | | -1.492 | 0.1357 | ?- |
| | | 17:75522194 | c | g | | MAC≤3 | -1.199 | 0.2306 | -? |
| | rs200434678 | 17:75522195 | a | g | MAC≤3 | | -2.035 | **0.04183** | ?- |
| | | 17:75522294 | a | g | MAC≤3 | | -0.752 | 0.4518 | ?- |
| | rs200228117 | 17:75523335 | a | g | MAC≤3 | MAC≤3 | -2.124 | **0.03368** | -- |
| | | 17:75524351 | a | g | | MAC≤3 | 1.243 | 0.214 | +? |
| ***LTA*** **(PAV 5%)** | rs2229092 | 6:31572980 | a | c | 0.03434 | 0.03556 | -3.789 | **0.0001475** | -- |

**Supplementary Table 7 – Replication of suggestive gene aggregate results**

| GENE | MAF-class | MetaSKAT P | T2D UKBB_Nephropathy_loose P | TOPMED P | FD GWAS P | THLBB P |
|---|---|---|---|---|---|---|
| *NAT16* | 0.1 | $1.4\times10^{-4}$ | 0.316296 | 0.5555 | 0.5813022 | NA |
| *LTA* | 0.05 | $1.5\times10^{-4}$ | NA | NA | NA | 0.4295364 |
| *SLC10A6* | 0.1 | $2.7\times10^{-4}$ | 0.162221 | 0.5622 | 0.8385422 | 0.7177943 |
| *TSEN54* | 0.01 | $3.7\times10^{-4}$ | 0.438022 | 0.7239 | 0.8519525 | 0.7194490 |

**Supplementary Table 8 – Lookup of monogenic kidney disease-causing genes on WES-WGS meta-analysis for DKD. A** meta-analysis p-values for all nominally (*P*<0.05) DKD-associated genes at MAF<0.1, MAF<0.05 and MAF<0.01, and **B** nominally significant genes highlighted among all the tested genes in different kidney disease categories.

| GENE | *P*(MAF=0.1) | *P*(MAF=0.05) | *P*(MAF=0.01) |
|---|---|---|---|
| **CYSTIC KIDNEY DISEASE OR NEPHRONOPHTHISIS** | | | |
| *CCDC28B* | 0.779 | 0.779 | **0.009** |
| *CEP104* | 0.221 | 0.196 | **0.007** |
| *CEP164* | 0.113 | 0.114 | **0.022** |
| *CSPP1* | 0.093 | **0.030** | 0.238 |
| *DCDC2* | **0.018** | **0.018** | 0.119 |
| *DDX59* | 0.686 | 0.686 | 0.680 |
| *HOXA4* | **0.0008** | **0.0008** | 0.545 |
| *IFT122* | **0.045** | **0.044** | 0.075 |
| *IQCB1* | **0.015** | **0.009** | 0.386 |
| *KIF14* | **0.002** | **0.002** | **0.026** |
| *UMOD* | 0.366 | 0.366 | **0.029** |
| **SYNDROMIC CAKUT** | | | |
| *ATXN10* | **0.026** | **0.026** | **0.026** |
| *BSCL2* | **0.023** | **0.023** | **0.023** |
| *EP300* | **0.048** | 0.078 | 0.332 |
| *ERCC8* | **0.026** | **0.026** | **0.026** |
| *FANCA* | 0.376 | 0.244 | **0.022** |
| *FAT4* | **0.041** | **0.032** | 0.384 |
| *HNRNPUL2-BSCL2* | **0.027** | **0.027** | **0.027** |
| *KCNH2* | 0.311 | 0.236 | **0.034** |
| *PAX8* | **0.021** | **0.021** | 1.000 |
| *POC1A* | **0.002** | **0.002** | **0.002** |
| *SALL4* | **0.035** | **0.035** | 0.524 |
| *WFS1* | 0.207 | 0.210 | **0.039** |
| **ISOLATED CAKUT** | | | |
| *DSTYK* | **0.024** | **0.038** | **0.026** |

| | | | |
|---|---|---|---|
| *SRGAP1* | 0.428 | 0.428 | **0.012** |
| **CHRONIC LOMERULONEPHRITIS** | | | |
| **RENAL TUBULOPATHIES** | | | |
| *SCN4A* | 0.413 | 0.413 | **0.037** |
| *SCNN1A* | **0.015** | **0.015** | 0.893 |
| *SCNN1G* | **0.042** | **0.042** | **0.042** |
| *WNK4* | **0.034** | 0.088 | 0.118 |
| **NEPHROLITHIASIS OR NEPHROCALCINOSIS** | | | |
| *CLDN16* | **0.015** | **0.015** | 0.150 |

**B**

**Cystic kidney disease or nephronophthisis** NPHP1 *INVS\NPHP2* *NPHP3* ***NPHP4*** ***IQCB1\NPHP5***
*CEP290\NPHP6* *GLIS2\NPHP7* *RPGRIP1L\NPHP8* *NEK8\NPHP9* *SDCCAG8\NPHP10*
*TMEM67\NPHP11* *TTC21B\NPHP12* *WDR19\NPHP13* *ZNF423\NPHP14* ***CEP164\NPHP15***
*ANKS6\NPHP16* *IFT172\NPHP17* *CCDC41\CEP83* ***DCDC2\NPHP19*** *MAPKBP1\NPHP20*
*IFT81\CDV-1* *TRAF3IP1\SLS9* *XPNPEP3\NPHPL1* *FAN1\MTMR15* *PKHD1\ARPKD*
*INPP5E\JBTS1* *TMEM216\JBTS2* *AHI1\JBTS3* *ARL13B\JBTS8* *CC2D2A\JBTS9*
*KIF7\JBTS12* *TCTN1\JBTS13* *TMEM237\JBTS14* *CEP41\TSGA14* *TMEM138\JBTS16*
*C5orf42\JBTS17* *TCTN3\JBTS18* *TMEM231\JBTS20* ***CSPP1\JBTS21*** *PDE6D\JBTS22*
*KIAA0586\JBTS23* *TCTN2\JBTS24* ***CEP104\JBTS25*** *KIAA0556\JBTS26* *MKS1\MKS1*
*B9D1\MKS9* *B9D2\MKS10* ***KIF14\MKS12*** *TMEM107\MKS13* *BBS1\BBS1* *BBS2\BBS2* *ARL6\BBS3*
*BBS4\BBS4* *BBS5\BBS5* *MKKS\BBS6* *BBS7\BBS7* *TTC8\BBS8* *PTHB1\BBS9* *BBS10\BBS10* *TRIM32\BBS11*
*BBS12\BBS12* *WDPCP\BBS15* *LZTFL1\BBS17* *BBIP1\BBS18* *IFT27\BBS20* *DDX59\OFD5* *SCLT1\OFD9*
*C2CD3\OFD14* *KIAA0753\OFD15* ***IFT122\CED1*** *WDR35\CED2* *IFT43\CED3*
*IFT80\SRTD2* *DYNC2H1\SRTD3* *NEK1\SRTD6* *WDR60\SRTD8* *IFT140\SRTD9* *WDR34\SRTD11*
*CEP120\SRTD13* *IFT57* *IFT52\SRTD* *ALMS1\ALMS* *PIK3R4\VPS15* *TXNDC15* *SLC41A1* *POC1B*
*HSD17B4\MFP2* *USH2A* ***UMOD*** ***HOXA4*** *HOXB6* *TBC1D32* **CCDC28B** EVC EVC2
*OFD1*

**Syndromic CAKUT** *B3GALTL* *BBS6* *BBS8* *BSCL2* *CD151* *CD96* *CEP290* *CHRNG* *CISD2*
*CTU2* *CYP21* *DACH1* *DHCR7* *EMG1* ***ERCC8*** *ESCO2* *ETFA* *ETFB* *ETFDH*
*FANCA* *FANCB* *FANCD2* *FANCE* *FANCI* *FANCL* ***FAT4*** *FOXP1* *HES7* *HYLS1*
*ICK* *IFT46* *IFT74* *INPP5E* *ITGA3* *JAM3* *LFNG* *LMNA* *LRIG2* *LRP2*
*LRP4* *MESP2* *MKS3* *PEX5* *PMM2* ***POC1A*** *PROK2* *RECQL4* *ROR2* *RPS19*

| | | | | | | | | | | |
|---|---|---|---|---|---|---|---|---|---|---|
| | *SCARF2* | *STRA6* | *TMCO1* | *TWIST2* | *UBR1* | *PEX1* | *PIGL* | *PIGO* | *PIGN* | *PIGT* |
| | *PIGV* | *PIGY* | *PTF1A* | ***WFS1*** | *WNT3* | *ZMPSTE24* | *ACTB* | *ACTG1* | *AIFM3* | ***ATXN10*** |
| | *BICC1* | *BMP7* | *BRAF* | *CDC5L* | *CREBBP* | *DACT1* | ***EP300*** | *ESRRG* | *FBN1* | *FGFR1* |
| | *FGFR3* | *FGF10* | *FGF8* | *FGF3* | *FMN1* | *FOXC1* | *FOXF1* | *GDF3* | *GDNF* | *GFRA1* |
| | *GLI2* | *HOXA13* | *HOXD13* | *JAG1* | *KAT6B* | *KCTD1* | ***KCNH2*** | *KRAS* | *LMX1B* | *LPP* |
| | *MAP2K1* | *MAP2K2* | *MLL2/ KMT2D* | | *MYCN* | *NFIX* | *NOTCH2* | ***PAX8*** | *PKD1* | *PKD2* |
| | *PROKR2* | *PTPN11* | *RAF1* | *RAI1* | ***SALL4*** | *SEMA3A* | *SEMA3E* | *SETBP1* | *SHH* | *SF3B4* |
| | *SNAP29* | *SOS1* | *SOX9* | *SRCAP* | *TBX1* | *TBX3* | *TFAP2A* | *TP63* | *TRPS1* | *TSC1* |
| | *TSC2* | *WNT5A* | *ARID1B* | *DIS3L2* | *FGFR2* | *GDF6* | *GLI3* | *PCSK5* | *PTEN* | *RPS24* |
| | *VANGL1* | *AXIN1* | *H19* | *KCNQ1OT1* | *NIPBL* | *CDKN1C* | *CHD7* | *AMER1* | *ATP7A* | *BCOR* |
| | *DLG3* | *FAM58A* | *FLNA* | *GPC3* | *MID1* | *NSDHL* | *OFD1* | *PIGA* | *PORCN* | *SMC1A* |
| | *UPF3B* | *ZIC3* | *GDF11* | *OSR1* | *TTC30A* | *UBE3A* | *SH2B1* | | | |
| **Isolated CAKUT** | | *ACE* | *AGT* | *AGTR1* | *CHRM3* | *ETV4* | *FRAS1* | *FREM1* | *FREM2* | *GRIP1* |
| | *HPSE2* | *ITGA8* | *REN* | *TRAP1* | *FGF20* | *BMP4* | *CHD1L* | *CRKL* | ***DSTYK*** | *EYA1* |
| | *GATA3* | *GREB1L* | *HNF1B* | *MUC1* | *NRIP1* | *PAX2* | *PBX1* | *RET* | *ROBO2* | *SALL1* |
| | *SIX1* | *SIX2* | *SIX5* | *SLIT2* | *SOX17* | ***SRGAP1*** | *TBX18* | *TNXB* | *UPK3A* | *WNT4* |
| | *KAL1* | | | | | | | | | |
| **Chronic glomerulonephritis** | | | *ADAMTS13* | *CFI* | *COL4A4* | *CFB* | *CFHR3* | *CFHR5* | *EIF2AK3* | *FN1* |
| | *FOXC2* | *GSN* | *LYZ* | *THBD* | *SPRY2* | *C3* | *CD46* | *CFH* | *CFHR1* | *COL4A3* |
| | *COL4A5* | *COL4A6* | | | | | | | | |
| **Renal tubulopathies** | | *ATP6B1* | *ATP6V1C2* | *BCS1L* | *BSND* | *COG6* | *COQ9* | *CYP11B1* | *CYP17A1* | *CYP27B1* |
| | *EGF* | *FGF23* | *GALNT3* | *HSD11B2* | *MRPS22* | *RRM2B* | ***SCNN1A*** | *SCNN1B* | *SLC12A3* | *SLC26A4* |
| | *SLC2A2* | *SLC4A4* | *SLC4A5* | *SLC6A19* | *SLC7A7* | *SUCLA2* | *TRPM6* | *VIPAR* | *VIPAS39* | *WNK1* |
| | ***WNK4*** | *AP2S1* | *AVP* | *CACNA1S* | *CNNM2* | *CUL3* | *EHHADH* | *FXYD6-FXYD2* | | *GNA11* |
| | *KCNJ5* | *KLHL3* | *NR3C2* | *SAC (ADCY10)* | | ***SCN4A*** | *CLCNKA* | *CLCKNB* | *SLC36A2* | *SLC6A20* |
| | *AQP2* | ***SCNN1G*** | *SLC5A2* | *AVPR2* | | | | | | |
| **Nephrolithiasis or nephrocalcinosis** | | | *AGXT* | *ALDOB* | *ALPL* | *APRT* | *ATP6V0A4* | *ATP6V1B1* | *ATP7B* | *CA2* |
| | *CLCNKB* | ***CLDN16*** | *CLDN19* | *CTNS* | *CYP24A1* | *FAH* | *FAM20A* | *G6PC* | *GRHPR* | *HOGA1* |
| | *KCNJ1* | *KCNJ10* | *SLC12A1* | *SLC26A1* | *SLC2A2* | *SLC34A3* | *XDH* | *HNF4A* | *SLC9A3R1* | *CASR* |
| | *SLC22A12* | *SLC2A9* | *SLC34A1* | *SLC3A1* | *SLC4A1* | *SLC7A9* | *VDR* | *CLCN5* | *HPRT1* | *OCRL* |
| Nephrotic syndrome | | *ADCK4* | *ALG1* | *APOA1* | *ARHGDIA* | *AVIL* | *CD2AP* | *COQ2* | *COQ6* | *CUBN* |
| | *CRB2* | *DGKE* | *EMP2* | *FAT1* | *ITGA3* | *ITGB4* | *KANK1* | *KANK2* | *KANK4* | *LAGE3* |

| | | | | | | | | | |
|---|---|---|---|---|---|---|---|---|---|
| *LAMB2* | *LCAT* | *MAGI2* | *MYO1E* | *NEU1* | *NPHS1* | *NPHS2* | *NUP107* | *NUP133* | *NUP205* |
| *NUP85* | *NUP93* | *OSGEP* | *PDSS2* | *PLCE1* | *PTPRO* | *SCARB2* | *SGPL1* | *SMARCAL1* | *TP53RK* |
| *TPRKB* | *TTR* | *VPS33B* | *WDR73* | *XPO5* | *ACTN4* | *ANLN* | *ARHGAP24* | *INF2* | *LMX1B* |
| *MYH9* | *PODXL* | *TRPC6* | *WT1* | *IKBKAP* | *NXF5* | *APOE* | *APOL1* | *GPC5* | *SYNPO* |

**Supplementary Table 9 –Tested variants in the associated sliding-window, promoter and enhancer associations**

| GENE | RSID | POSITION | ALLELE1 | ALLELE2 | MAC | BETA* | FIRTH P-VALUE |
|---|---|---|---|---|---|---|---|
| **SLIDING-WINDOW 12Q14.3** | rs575374987 | 12:108104058 | C | T | ≤3 | -0.45785 | 0.829189 |
| | | 12:108104432 | C | A | ≤3 | -1.73385 | 0.415624 |
| | rs561150045 | 12:108104469 | G | A | ≤3 | -0.515692 | 0.807619 |
| | rs190528531 | 12:108105375 | G | A | ≤3 | -1.87318 | 0.376391 |
| | rs74676695 | 12:108106003 | G | A | ≤3 | -1.87063 | 0.377378 |
| | rs78452498 | 12:108106081 | C | A | 4 | -2.34618 | 0.147223 |
| | | 12:108106137 | TTAATA A | T | ≤3 | -1.76798 | 0.377696 |
| | rs531800089 | 12:108106452 | C | A | ≤3 | -1.88274 | 0.373646 |
| | | 12:108106913 | C | T | ≤3 | -0.506038 | 0.811192 |
| | | 12:108106970 | C | T | ≤3 | -2.25076 | 0.236695 |
| | | 12:108107279 | G | C | ≤3 | -1.98858 | 0.347517 |
| | rs192506403 | 12:108107522 | C | T | ≤3 | -1.1887 | 0.494028 |
| | | 12:108107725 | T | C | ≤3 | -2.05204 | 0.333586 |
| | rs117656942 | 12:108107793 | C | T | ≤3 | -1.74338 | 0.383098 |
| **SLIDING-WINDOW 4Q22.3** | rs151208036 | 4:75122307 | G | A | ≤3 | 0.880516 | 0.678125 |
| | rs376478683 | 4:75122408 | AGAAG | A | ≤3 | 0.578048 | 0.785264 |
| | | 4:75122522 | GA | G | ≤3 | -1.98729 | 0.348375 |
| | | 4:75122526 | A | G | ≤3 | -1.85719 | 0.380384 |
| | | 4:75122863 | G | A | 4 | 2.56836 | 0.113218 |
| | | 4:75123692 | GCTTT | G | ≤3 | 0.971009 | 0.610347 |
| | | 4:75123736 | CA | C | ≤3 | 0.656683 | 0.756079 |
| | | 4:75123739 | A | C | ≤3 | 0.658244 | 0.755548 |
| | | 4:75123742 | G | T | ≤3 | -0.811792 | 0.513137 |
| | rs72662456 | 4:75124048 | C | T | ≤3 | 2.08135 | 0.326227 |
| | | 4:75124093 | C | T | ≤3 | 0.693639 | 0.625782 |

| | | | | | | | |
|---|---|---|---|---|---|---|---|
| | | 4:75124193 | A | G | ≤3 | 1.40742 | 0.419141 |
| | rs114761270 | 4:75124427 | C | T | 7 | 2.71697 | 0.0397647 |
| | rs528483191 | 4:75124599 | CTA | C | 4 | -0.29815 | 0.774696 |
| | rs140007606 | 4:75124646 | C | T | 6 | 1.90913 | 0.0584707 |
| | rs747665362 | 4:75124654 | C | G | ≤3 | 2.2326 | 0.292939 |
| | rs770601133 | 4:75124950 | T | G | 10 | -0.379054 | 0.582752 |
| | | 4:75125317 | A | C | ≤3 | 1.87794 | 0.343649 |
| | | 4:75125568 | C | T | ≤3 | -0.515692 | 0.807619 |
| | rs147887794 | 4:75125585 | A | C | ≤3 | 0.979194 | 0.607488 |
| | rs62314936 | 4:75125980 | T | A | 7 | 1.61021 | 0.100681 |
| **PROMOTER 9Q21.11 (*CTSL3P*)** | | 9:87792152 | C | CAG | 40 | -1.09871 | 0.0033829 |
| | rs141341525 | 9:87792611 | G | A | 5 | 0.318125 | 0.738099 |
| | rs12349827 | 9:87792755 | G | T | 8 | -2.04777 | 0.0297324 |
| | | 9:87793023 | G | C | ≤3 | -0.552815 | 0.794438 |
| | rs12349926 | 9:87793063 | G | A | 23 | -1.96123 | 0.000608268 |
| | rs566499267 | 9:87793119 | A | G | ≤3 | 1.82619 | 0.355895 |
| **PROMOTER 2Q14.2 (*LINC01794*)** | rs191082134 | 2:40755328 | C | T | ≤3 | -1.77203 | 0.403132 |
| | | 2:40755562 | A | T | ≤3 | -0.514951 | 0.808033 |
| | | 2:40755569 | G | C | ≤3 | -2.02071 | 0.340118 |
| | rs183413211 | 2:40755782 | T | A | 6 | -2.91437 | 0.0361816 |
| | | 2:40756209 | TA | T | 4 | -2.72118 | 0.0681172 |
| **ENHANCER 18Q12.3 (METTL4)** | rs530496044 | 18:2351117 | A | G | ≤3 | 1.98211 | 0.34921 |
| | rs16943099 | 18:2351193 | G | C | 15 | -2.08034 | 0.000208644 |
| **ENHANCER 9Q21.11 (*CTSL*)** | | 9:87793023 | G | C | ≤3 | -0.552815 | 0.794438 |
| | rs12349926 | 9:87793063 | G | A | 23 | -1.96123 | 0.000608268 |
| | rs566499267 | 9:87793119 | A | G | ≤3 | 1.82619 | 0.355895 |
| | rs549934850 | 9:87793172 | G | A | ≤3 | -0.570487 | 0.787453 |

* BETA: effect size estimate for ALLELE1

### Supplementary Table 10 - FinnDiane Study group

Heinola Health Center: P.Hentunen, J.Lagerstam
Helsinki University Central Hospital (HUS): Department of Medicine, Division of Nephrology A.Ahola, M.Feodoroff, D.Gordin, O.Heikkilä, K.Hietala, M.Korolainen J.Kytö, S.Lindh, K.Pettersson-Fernholm, A.Sandelin, L.Thorn, J.Tuomikangas, T.Vesisenaho, J.Wadén
Herttoniemi Hospital, Helsinki: V. Sipilä
Hospital of Lounais-Häme, Forssa: T. Kalliomäki, J. Koskelainen, R. Nikkanen, N.Savolainen, H.Sulonen, E.Valtonen
Hyvinkää Hospital: L. Norvio, A. Hämäläinen
Iisalmi Hospital: E.Toivanen
Jokilaakso Hospital, Jämsä: A.Parta, I.Pirttiniemi
Jorvi Hospital, HUS: S. Aranko, S.Ervasti, R.Kauppinen-Mäkelin, L. Pekkonen, A. Kuusisto, T.Leppälä, K. Nikkilä
Jyväskylä Health Center, Kyllö: K. Nuorva, M.Tiihonen
Kainuu Central Hospital, Kajaani: S. Jokelainen, K. Kananen, M.Karjalainen, P.Kemppainen, A-M.Mankinen, A.Reponen, M.Sankari
Kerava Health Center: H.Stuckey, P.Suominen
Kirkkonummi Health Center: A.Lappalainen, M.Liimatainen, J.Santaholma
Kivelä Hospital, Helsinki: A.Aimolahti, E.Huovinen
Koskela Hospital, Helsinki: V.Ilkka, M.Lehtimäki
Kotka Health Center: E.Pälikkö-Kontinen, A.Vanhanen
Kouvola Health Center: E.Koskinen, T.Siitonen
Kuopio University Hospital: E.Huttunen, R.Ikäheimo, P.Karhapää, P.Kekäläinen, M.Laakso, T.Lakka, E.Lampainen, L.Moilanen, L.Niskanen, U.Tuovinen, I.Vauhkonen, E.Voutilainen
Kuusamo Health Center: T.Kääriäinen, E.Isopoussu
Kuusankoski Hospital: E.Kilkki, I.Koskinen, L.Riihelä
Laakso Hospital, Helsinki: T.Meriläinen, P.Poukka, R.Savolainen, N.Uhlenius
Lahti City Hospital: A.Mäkelä, M.Tanner Lapland Central Hospital, Rovaniemi: L.Hyvärinen, K.Lampela, S.Pöykkö, T.Rompasaari, S.Severinkangas, T.Tulokas
Lappeenranta Health Center: P. Erola, L. Härkönen, P.Linkola, I.Pulli, E.Repo
Lohja Hospital T.Granlund, K.Hietanen, M.Porrassalmi, M.Saari, T.Salonen, M.Tiikkainen,
Länsi-Uusimaa Hospital, Tammisaari: I.-M.Jousmaa, J.Rinne
Loimaa Health Center: A. Mäkelä, P. Eloranta
Malmi Hospital, Helsinki: H.Lanki, S.Moilanen, M.Tilly-Kiesi
Mikkeli Central Hospital: A.Gynther, R.Manninen, P.Nironen, M.Salminen, T.Vänttinen
Mänttä Regional Hospital: I.Pirttiniemi, A-M.Hänninen
North Karelian Hospital, Joensuu: U-M.Henttula, P.Kekäläinen, M.Pietarinen, A.Rissanen, M.Voutilainen
Nurmijärvi Health Center: A.Burgos, K.Urtamo
Oulaskangas Hospital, Oulainen: E.Jokelainen, P-L.Jylkkä, E.Kaarlela, J.Vuolaspuro
Oulu Health Center: L.Hiltunen, R.Häkkinen, S.Keinänen-Kiukaanniemi
Oulu University Hospital: R.Ikäheimo
Päijät-Häme Central Hospital: H.Haapamäki, A.Helanterä, S.Hämäläinen, V.Ilvesmäki, H.Miettinen
Palokka Health Center: P.Sopanen, L.Welling

Pieksämäki Hospital: V.Sevtsenko, M.Tamminen
Pietarsaari Hospital: M-L.Holmbäck, B.Isomaa, L.Sarelin
Pori City Hospital: P.Ahonen, P.Merisalo, E.Muurinen, K.Sävelä
Porvoo Hospital: M.Kallio, B.Rask, S.Rämö
Raahe Hospital: A.Holma, M.Honkala, A.Tuomivaara, R.Vainionpää
Rauma Hospital: K.Laine, K.Saarinen, T.Salminen
Riihimäki Hospital: P.Aalto, E.Immonen, L.Juurinen
Salo Hospital: A.Alanko, J.Lapinleimu, P.Rautio, M.Virtanen
